# A Descriptive Analysis of Calls to the NSW Teratogen Information Service regarding use of anti-infectives during pregnancy

**DOI:** 10.1101/2022.06.22.22276740

**Authors:** Helen Ritchie, Elizabeth Hegedus, Joanne Ma, Debra Kennedy

## Abstract

**Background:** MotherSafe is a free telephone-based counseling service for Australian consumers and health-care providers concerned about drug exposures during pregnancy and breastfeeding. Anti-infectives are the most commonly prescribed drugs for pregnant women. This study aims to provide a retrospective, descriptive analysis of prospectively collected calls received by MotherSafe regarding anti-infective exposures during pregnancy between 2000 and 2020. Aggregate data were examined by type of caller, reason for call, pregnancy category and exposure type. Inductive thematic analysis of the comments recorded by MotherSafe counsellors at the time of call was undertaken.

**Results:** Over the study period, 25,890 calls related to exposure to anti-infectives during pregnancy (antibiotic, antiviral, and antifungal medications). Calls from patients were dominated by low-risk exposures (pregnancy category A) to drugs while calls from health care professionals related to drugs with limited human information (pregnancy category B3). Analysis of MotherSafe counsellor comments revealed over 200 instances of concerns relating to health care professional advice to the patient. Three themes emerged: incorrect or conflicting advice, poor counselling, and refusal to treat, prescribe or dispense. It is likely that these comments are biased to the negative as patients would not call MotherSafe if they were happy with HCP advice. However, the findings are concerning as they reveal an underlying lack of knowledge in some health care professionals which may have led to undertreatment of patients. This study reinforced the importance of Teratogen Information Services such as MotherSafe in providing counselling and clear communication of evidence-based information to guide decision-making, reducing potential emotional distress in pregnant women, and optimizing maternal, pregnancy and infant outcomes.

## Introduction

Up to 93% of women in first world countries are estimated to use prescription drugs at some time during pregnancy [1]. Concerns about adverse outcomes of medication use during pregnancy can induce anxiety, leading to women seeking information prior to or during gestation [2].

Low health literacy has been significantly associated with increased perception of risk regarding medication use during pregnancy [3]. While the underestimation of teratogenic risks could lead to congenital malformations or fetotoxicity, the overestimation of risk could result in non-adherence to treatment, undertreatment of serious maternal medical conditions or increased tendency to abort [4, 5]. It is therefore important that risk estimates of medication exposures during gestation are clearly conveyed in a format that promotes appropriate medicine use.

Drug information resources available to Australian health professionals include Obstetric Drug Information services such as MotherSafe in New South Wales (NSW), NPS MedicineWise, the Prescribing Medicines in Pregnancy database maintained by the Therapeutic Goods Administration (TGA), Australian Therapeutic Guidelines and the Australian Medicines Handbook. Prescription and over-the counter medicines are classified into non-hierarchical categories (A, B1-3, C, D and X) by the TGA based on risk level associated with gestational drug use, as determined by the quality and quantity of data available (Table 1).

**Table 1.**
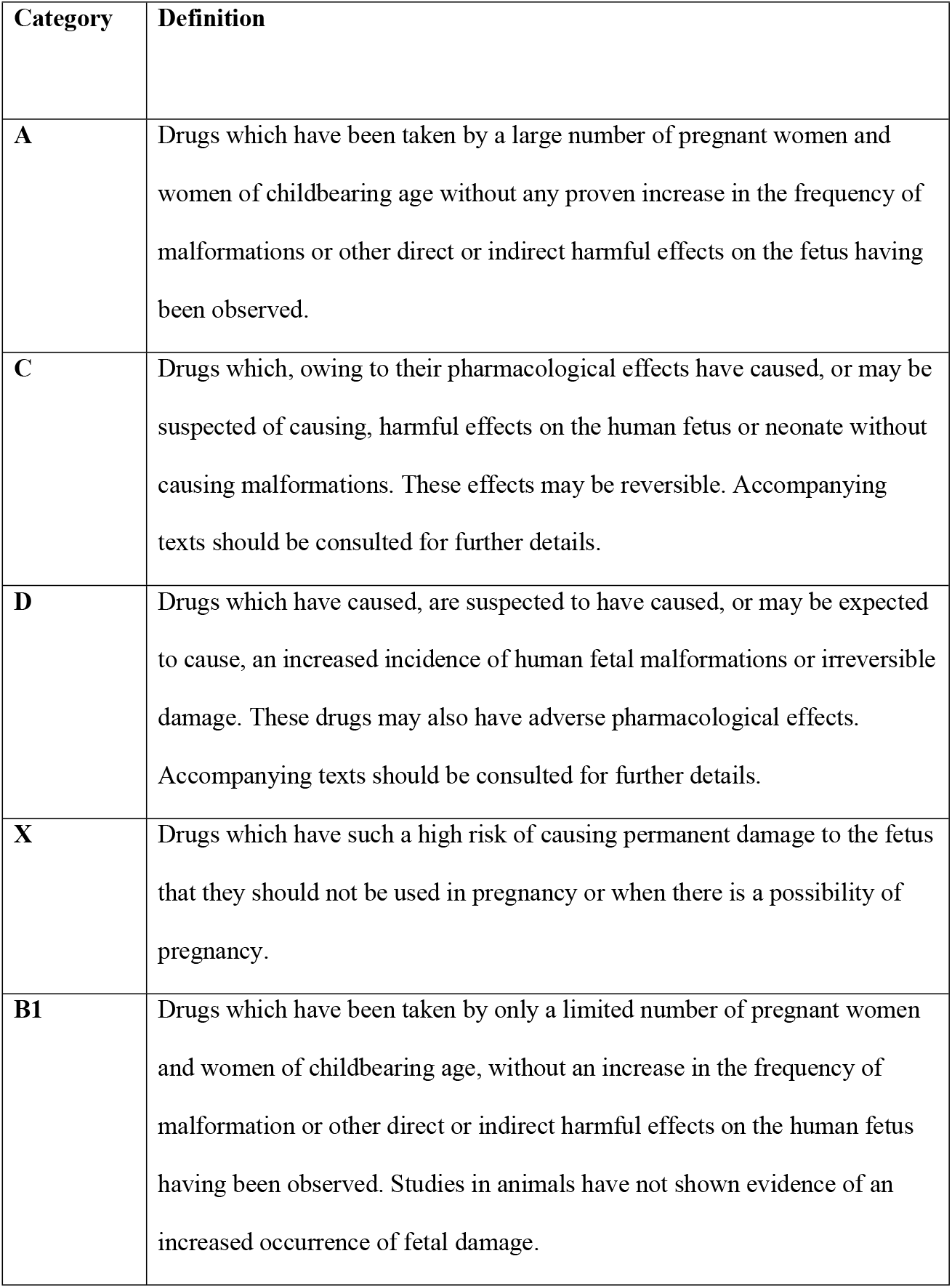
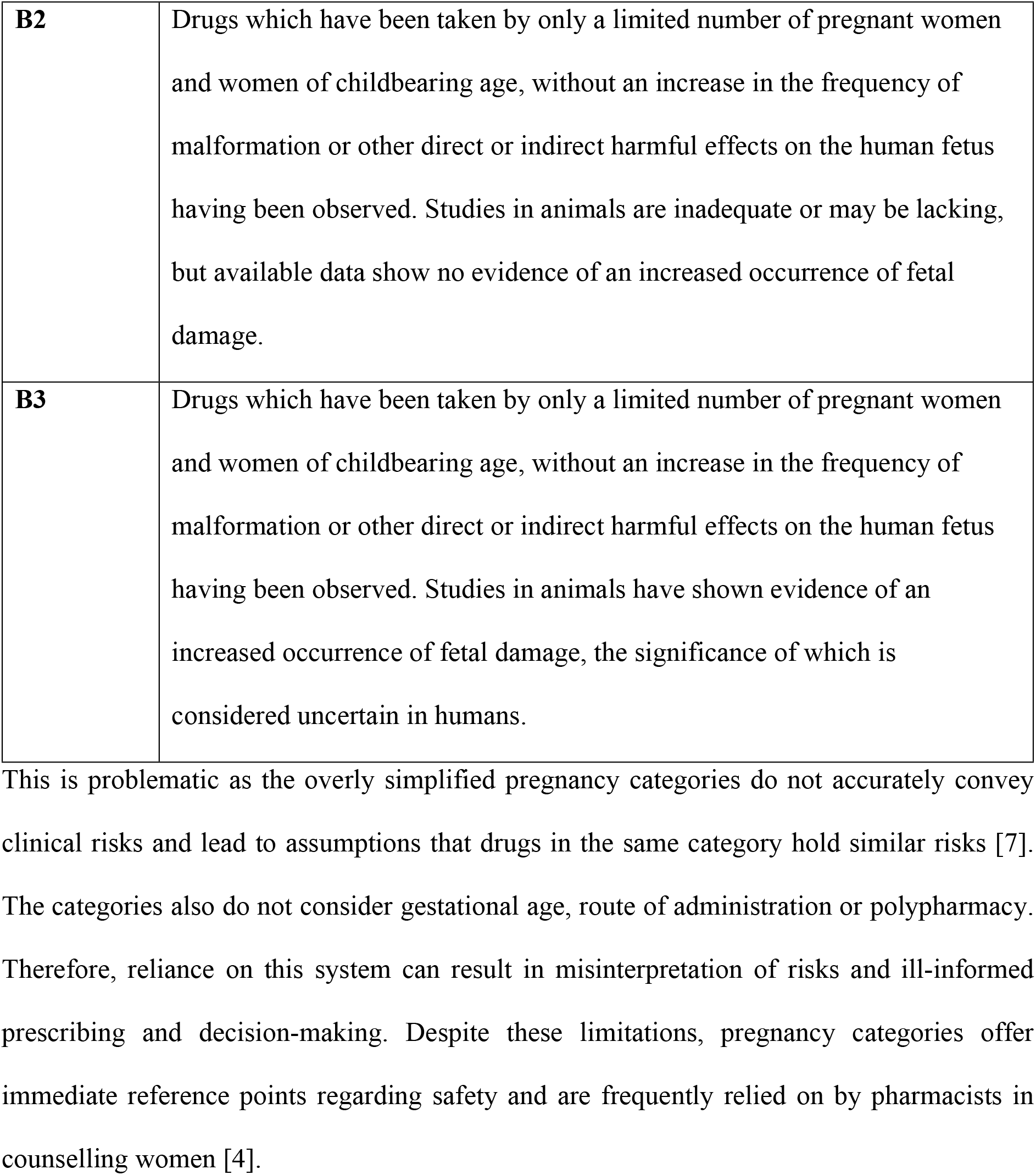
Australian categorisation system for registered medicines in pregnancy TGA [6].

Counselling and clear communication of benefits associated with medication use (and risks of an untreated maternal medical condition) may reduce perceived risks and improve decision-making. Twenty-four percent of pregnant Australians seek information from drug information centres, such as MotherSafe [8]. MotherSafe is a Teratogen Information Service based at the Royal Hospital for Women in New South Sales (NSW), that provides evidence-based information and counselling for women and healthcare professionals (HCPs) concerned about risks of various exposures (including medication) during pregnancy and breastfeeding. Anti-infectives are the most commonly prescribed drugs for pregnant women (39.8%) [9]. This study aims to provide a retrospective, descriptive analysis of calls received by MotherSafe regarding anti-infective exposures during pregnancy between 2000 and 2020.

## Materials and Methods

A retrospective descriptive analysis of collated call data from MotherSafe for the period January 2000 to December 2020 was conducted.

All data from MotherSafe calls are logged as they are received. Advice and counselling are provided regarding exposures during pregnancy and breastfeeding and include prescription drugs, over-the-counter (OTC) and off-the-shelf medications as well as recreational drugs, infections, vaccines and occupational exposures.

Information collected from each phone call includes: date of call, caller type (patient, family or friend, HCPs), residential location (postcode), maternal age, reason for call (pregnant, planning pregnancy, breastfeeding), gestational age, and exposures of concern. Additional comments are also sometimes added by the MotherSafe counsellor during the call. Exposures were classified by MotherSafe counsellors into one of 40 categories based on their general class. If a caller enquires about several potential exposures, these are separately listed. Therefore, the number of enquiries is greater than the number of calls. For this analysis, all data from calls relating to anti-infective (antibiotic, antifungal, and antiviral) exposures during pregnancy were analyzed. Data was first reviewed for spelling mistakes and input errors during data entry. The aggregated dataset was analyzed by enquiry for year, maternal and gestational ages, type, and location of caller.

Inductive thematic analysis of the comments recorded by MotherSafe counsellors at the time of call was undertaken using NVivo software Version 11.0 (QSR International). An initial scan of the data was performed to identify mis-coded data. Codes were then generated for the entire transcript set by two researchers and results were compared. Iterative comparison and refinement resulted in a final coding structure which was then examined to explore patterns and relationships between codes and concepts [10].

### Statistics

Statistical analysis was performed using IBM SPSS Statistics (version 26). A chi-square test of independence was conducted between reason for call and anti-infective exposure. Statistical significance was *p* < 0.05. Cramer’s V was calculated to estimate the strength of association.

All data was provided fully anonymized before analysis so informed consent was not required by ethics committee. The study was approved by the South-East Sydney Illawarra Area Health Service’s Human Research Ethics Committee (reference number 07/131).

## Results

Between January 2000 and December 2020, MotherSafe received 333,820 telephone enquiries regarding exposures during pregnancy and/or breastfeeding. Of these, 25,890 calls related to exposure to anti-infectives during pregnancy (including termination of pregnancy, and pregnancy whilst breastfeeding). The majority of callers were women calling for themselves (74.5%) with HCP calls comprising most of the remainder. Of these, the majority were general practitioners (60%). Over the 21-year period of this study, only 2.8% of calls were from family or friends and these were excluded from further statistical analysis.

Regardless of caller type, over half the calls related to antibiotic exposure with the calls relating to antivirals and antifungals equally divided (Table 2). While the difference was statistically significant (*X*^2^ = 350.4, *p* < 0.005), the association was small [11]Cramer’s V = 0.109). The mean age of the callers (32.4 years) was slightly higher than the Australian average of 31.9 years for women giving birth in 2018 [12]. Breastfeeding callers were significantly older than pregnant callers.

**Table 2:** Number of enquiries to MotherSafe (2000-2020) regarding anti-infective exposures during pregnancy by year and reason for call, maternal age, and gestational age.

|  | Pregnancy | Pregnancy & Breastfeeding | Termination of Pregnancy | Total (percent) |
| --- | --- | --- | --- | --- |
| Antibiotic Exposures | 14406 | 423 | 114 | 14,943 (55.9%) |
| Antifungal Exposures | 5777 | 114 | 24 | 5,915 (22.1%) |
| Antiviral Exposures | 5795 | 87 | 11 | 5,893 (22.0%) |
| Mean ~~maternal~~ age<br>(years ± *SD*) | 32.4 ± 4.85 | 33.2 ± 4.53* | 31.9 ± 4.98 | |
| Mean gestational age<br>(weeks ± *SD*) | 20.2 ± 10.65 | 20.5 ± 13.64 | 7.6 ± 3.82* | |

\* Compared to pregnancy, $p < 0.05$
Note: there may be overlap in exposure enquiries per call (i.e. one call may involve enquiries regarding more than one exposure group).

Gestational age was recorded for 24,092 of calls. The average gestational age was 20 weeks for pregnant or pregnant and breastfeeding women. Women calling regarding termination of pregnancy called significantly earlier in pregnancy (7.6 weeks). Enquiries in the first trimester accounted for 36.0%, second trimester (24.6%) and third trimester (39.4%) of calls. In total, only 19.4% occurred during gestation weeks 2-8 (period of organogenesis).

The effect of drug categorization on call frequency was also examined. Adequate information was available to categorize 84% of drug enquiries, the remainder being too generic to classify e.g. ‘antibiotic’. There was a statistically significant association between caller type and pregnancy category (*X*^2^ = 2698.8, *p* < 0.005). The association was moderate [11] Cramer’s V = 0.277). Patients made more calls regarding pregnancy category A drugs while calls from HCPs were predominantly pregnancy category B3 (Fig 1).

**Fig 1:**
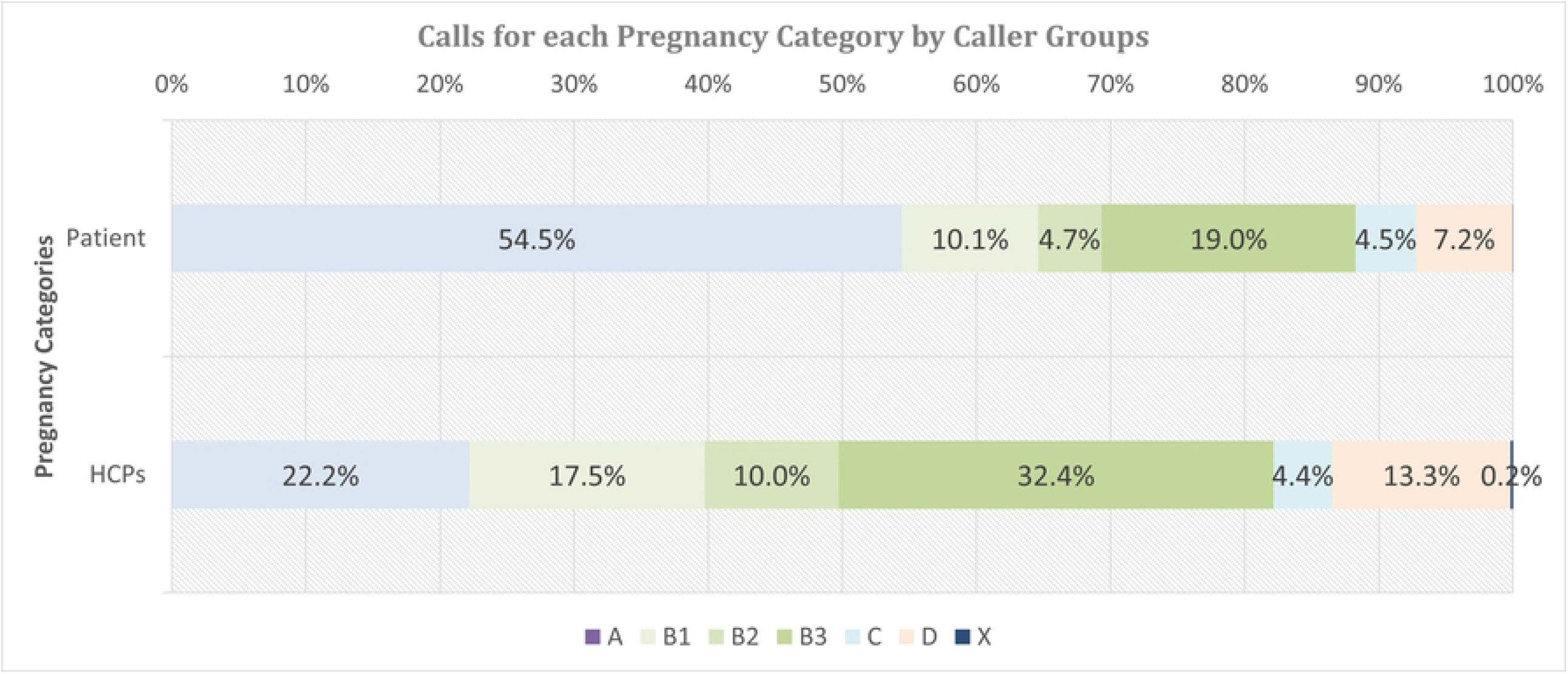
Pregnancy categories of all enquiries during pregnancy by caller type (patient or HCP)

The top ranked calls for individual drugs differed between patients and HCPs (listed in Table 3). Several drugs on the list are available topically (aciclovir, clotrimazole, metronidazole) or can be purchased without prescription (aciclovir, clotrimazole, fluconazole). Calls from patients were predominantly regarding pregnancy category A drugs such as the antibiotics, amoxicillin and cefalexin, and the topical, antifungal clotrimazole. Calls regarding the topical antiviral, aciclovir were also common. The most common reasons for calls from GPs related to the prescription-only medication, valaciclovir (pregnancy category B3). Pharmacists were most likely to call regarding the over-the counter, topical medication, aciclovir (pregnancy category B3) and the pharmacist-only oral medication, fluconazole (pregnancy category D).

**Table 3:** Top ranked enquiries by caller regarding anti-infective drugs with pregnancy category and prevalence

| Anti-infective | Pregnancy category | Prevalence among |  |  |
| --- | --- | --- | --- | --- |
|  |  | MotherSafe callers |  |  |
|  |  | Patients (%) | GP (%) | Pharmacists (%) |
| Amoxicillin | A | 16.1 | 2.2 | 1.0 |
| Clotrimazole | A | 14.8 | 4.1 | 7.3 |
| Aciclovir | B3 | 12.4 | 7.4 | 19.8 |
| Cefalexin | A | 9.9 | 1.5 | 0.6 |
| Fluconazole | D | 6.0 | 4.6 | 11.1 |
| Valaciclovir | B3 | 5.6 | 12.2 | 5.3 |
| Erythromycin | A | 4.2 | 2.3 | 0.8 |
| Metronidazole | B2 | 2.5 | 8.4 | 4.2 |
| Famciclovir | B1 | 2.2 | 2.7 | 7.1 |

## Analysis of Comments made to MotherSafe counselors regarding HCP advice

There were over 8000 comments (8361) recorded by MotherSafe counsellors across all caller types. The majority alluded to referrals for disease diagnosis and reassurances and advice given. However, there were 201 comments made by callers to MotherSafe counsellors relating to concerns regarding HCP advice. Three themes were identified from analysis of these comments (Table 4).

**Table 4:** Number of comments by patients regarding HCP advice by theme

| **Comment** | **GP or specialist** | **Pharmacist** | **Dentist** |
| --- | --- | --- | --- |
| Incorrect advice: conflicting with other HCP, HCP does not know answer, or incorrect advice given | 54<br>(54%) | 36<br>(36%) | 10<br>(10%) |
| Poor counselling: Advised not to take, overly cautious, or not reassuring | 13<br>(50%) | 11<br>(42%) | 2<br>(8%) |
| Patient untreated or undertreated due to refusal to treat, prescribe or dispense | 21<br>(31%) | 38<br>(56%) | 9<br>(13%) |

### Theme 1: Conflicting advice, lack of knowledge or incorrect advice by HCP

There were 100 calls where inadequate or incorrect advice was reportedly given by HCPs, stemming from a lack of knowledge, incorrect knowledge, or uncertainty regarding safety of the medication or treatment. This led to conflicting advice between HCPs and occurred at all stages of the treatment process, from prescription to dispensing. It includes HCPs stating that the patient cannot take anything during pregnancy or prescribing medication but also cautioning against using the medication.

Pharmacists also provided incorrect advice, particularly regarding the use of topical products such as aciclovir. Conflicting advice was particularly likely concerning drugs categorized as B3 or D. For example, a patient 34 weeks gestation was prescribed valaciclovir. However, before dispensing a pharmacist warned about possible deformities associated with the category B3 medication.

### Theme 2: Poor counselling: overly cautious or not reassuring

There were many calls from patients who were told by pharmacists that they “cannot use”, “shouldn’t use” or it was “not safe to use” the products during pregnancy. This is problematic for commonly used drugs such as topical aciclovir and clotrimazole that can be purchased OTC. Poor counselling also led to poor compliance with Tamiflu as neither GP nor pharmacist were reassuring.

### Theme 3: Refusal to treat, prescribe or dispense

There were several calls where an HCP had either refused to treat, prescribe or dispense a medication resulting in inadequate treatment particularly with pregnancy category A and B3 drugs. Examples include a patient with a parasitic infection whose doctor was reported to have said that the patient should be treated with metronidazole, but this was not possible as she was pregnant. Patients also encountered similar problems with dentists who refused to prescribe antibiotics for dental infections. Refusals were most common among pharmacists who would not dispense medication, including many involving topicals or categories where increased risks were perceived (B3 and D) e.g., aciclovir and fluconazole. While some pharmacists who refused to dispense did offer alternative treatment for example, topical clotrimazole instead of fluconazole many cases were simply blunt refusals.

## Discussion

Anti-infectives are the most commonly prescribed medication to pregnant women (Andrade et al., 2004). Almost a quarter of calls to MotherSafe were from HCPs. This large proportion of calls could either be attributed to their familiarity with TIS or their greater awareness of potential risks associated with some medications. Findings of this study support the latter as HCPs tended to call for information regarding exposure to medications with more restrictive pregnancy categories (Table 2). As pregnant women are excluded from drug clinical trials and the extrapolation of animal data to humans is complex, HCPs need to carefully weigh the risks and benefits based on available knowledge prior to prescribing, dispensing, or administering of medications. Overestimation of risks is not limited to pregnant women as HCPs also perceive increased risks, possibly due to scientific uncertainty and underuse of specialized resources [13]. The role of TIS such as MotherSafe have been instrumental in increasing confidence in prescribing and reducing overestimated teratogenic risks among HCPs [13, 14] potentially leading to better patient outcomes [15].

GPs and pharmacists are typically the first and last lines of contact as they oversee prescription and dispensing. They are the primary source of information in relation to pregnancy concerns. GPs, obstetricians and other specialists were an information source for 74% of pregnant women followed by the internet (60%) and pharmacists (54%) [8]. However, some pharmacists can be overly cautious in the provision of advice, especially for categories other than pregnancy category A, due to fear of liability [4]. Pharmacists may be the last point of contact prior to drug exposure, and possibly the only point of contact with a HCP in the case of OTC medication [16]. This places pharmacists in an important and influential role but this study found that, unfortunately, this influence on patients is not always beneficial. In our examination of comments made to MotherSafe counsellors, pharmacists were responsible for significant instances of potential undertreatment of patients due to overly cautious advice or refusing to dispense (Table 4).

No expert knowledge is required to post information online and there is no curation or explanation of claims and statements yet many pregnant women rely on this source for information [8]. Therefore reliable, evidence-based data can be overwhelmed or displaced. This can result in confusion, unnecessary stress, and fear amongst pregnant and breastfeeding women. Despite the decreasing awareness of the thalidomide disaster, its aftermath has been an increasing skepticism of medication safety in pregnancy [17]. A fear of birth defects was found to be the leading fear of women taking medication during pregnancy and is a possible cause of non-adherence and highlights the need for stronger communication of risks and benefits [18]. The frequency of patient calls regarding pregnancy category A drugs suggests this communication may be lacking.

While it is reassuring to note that there are many calls within the first trimester, patient enquiries continued into the second and third trimesters. Antifungal enquiries particularly increased during the third trimester as might be expected. Candida infections, or thrush, are experienced by up to 40% of women during pregnancy [19]. More than half the enquiries relating to fluconazole exposure (a commonly prescribed topical medication to treat thrush) occurred during the third trimester. Fluconazole is pregnancy category D whether it is at low dose to treat thrush or higher doses to treat meningitis. The large number of calls relating to this medication possibly is the result of a misunderstanding of the pregnancy categories again highlighting the shortcomings of pregnancy categorization e.g. dose, route of exposure and stage of pregnancy are rarely taken into account. This is supported by the analysis of comments with many instances of pharmacists reportedly refusing to dispense fluconazole to patients in their third trimester. Most topical medications would be expected to have limited systemic absorption yet frequently have the same pregnancy category as oral or intravenous routes of administration. This misclassification has become a common cause for concern by HCPs despite minimal risks. For example, the antiviral, aciclovir, can be purchased OTC and is categorized B3 but when used topically to treat cold sores, it is considered safe to use in pregnancy (Australian Medicines Handbook 2020). Yet aciclovir accounted for nearly 20% of calls from pharmacists.

It is clear from the number of calls by HCPs as well as the comments to MotherSafe that there is a need for improved access to accurate and up-to-date information by HCPs. Current resources for HCPs in Australia include MIMS, Australian Therapeutic Guidelines, Australian Medicines Handbook and the TGA’s Medicines in Pregnancy online database. However, an analysis of pregnancy information from the product information and simplified, consumer medicine information found them to be overly cautious, legalistic and risk averse [20].

Specialist databases such as Reprotox and TERIS are not freely available. Resources such as the Australian Therapeutic Guidelines and Prescribing medicines in Pregnancy Database only provide the pregnancy category with no explanation to assist counselling. The AMH avoids categorizations and provides only a compatible/incompatible guideline, again with no explanation which some HCPs have found unhelpful [4]. This is inadequate given the nuanced nature of counselling.

## Conclusion

The risks and benefits of taking a medication during pregnancy should be carefully assessed prior to it being prescribed, dispensed and administered by HCPs. Accurate, evidence-based information is essential for this decision-making to avoid preventable or adverse outcomes. Misinterpretation of risks is exacerbated by the current pregnancy categorization system which is not nuanced enough to support appropriate counselling. Overestimation of risks and fear of adverse outcomes unfavorably outweigh the perceived benefits resulting in increased hesitancy towards the commencement or adherence to medication. This was clear from the calls to MotherSafe, where HCPs would not prescribe or dispense medicine, and patients were hesitant to take prescribed medicine. Hence TISs such as MotherSafe play an important role in providing counselling and clear communication of evidence-based information to guide decision-making, reducing the potential emotional distress, and optimizing maternal, pregnancy and infant outcomes.

## Limitations of this study

The calls to MotherSafe may represent a biased sample as no educational, professional or background information was gathered. The calls are also largely limited to women of English-speaking background. The calls may not be a true representation of the encounters between HCPs and patients as the description of the events are those relayed by the caller and may be biased. However, it should be acknowledged that call represented the patient perception of the HCP advice. Finally, calls are likely to be biased as patients may be less inclined to call if they are happy with advice received by their HCP.

## Data Availability

Data cannot be shared publicly personal information may be revealed. Data are available from the South-East Sydney Illawarra Area Health Service's Human Research Ethics Committee (contact via Debra Kennedy) for researchers who meet the criteria for access to confidential data.

## Acknowledgments

We wish to acknowledge the ongoing support of the Royal Hospital for Women Foundation.

